# Consistent Multi-Omic Relationships Uncover Molecular Basis of Pediatric Asthma IgE Regulation

**DOI:** 10.1101/2024.06.05.24308502

**Authors:** Tara Eicher, Rachel S. Kelly, John Braisted, Jalal K. Siddiqui, Juan Celedón, Clary Clish, Robert Gerszten, Scott T. Weiss, Michael McGeachie, Raghu Machiraju, Jessica Lasky-Su, Ewy A. Mathé

## Abstract

Serum total immunoglobulin E levels (total IgE) capture the state of the immune system in relation to allergic sensitization. High levels are associated with airway obstruction and poor clinical outcomes in pediatric asthma. Inconsistent patient response to anti-IgE therapies motivates discovery of molecular mechanisms underlying serum IgE level differences in children with asthma. To uncover these mechanisms using complementary metabolomic and transcriptomic data, abundance levels of 529 named metabolites and expression levels of 22,772 genes were measured among children with asthma in the Childhood Asthma Management Program (CAMP, N=564) and the Genetic Epidemiology of Asthma in Costa Rica Study (GACRS, N=309) via the TOPMed initiative. Gene-metabolite associations dependent on IgE were identified within each cohort using multivariate linear models and were interpreted in a biochemical context using network topology, pathway and chemical enrichment, and representation within reactions. A total of 1,617 total IgE-dependent gene-metabolite associations from GACRS and 29,885 from CAMP met significance cutoffs. Of these, glycine and guanidinoacetic acid (GAA) were associated with the most genes in both cohorts, and the associations represented reactions central to glycine, serine, and threonine metabolism and arginine and proline metabolism. Pathway and chemical enrichment analysis further highlighted additional related pathways of interest. The results of this study suggest that GAA may modulate total IgE levels in two independent pediatric asthma cohorts with different characteristics, supporting the use of L-Arginine as a potential therapeutic for asthma exacerbation. Other potentially new targetable pathways are also uncovered.

## Introduction

Immunoglobulin E (IgE) is an antibody that binds to mast cells and basophils in response to allergen exposure, contributing to asthma exacerbation in children ^1^. Exacerbations result in episodes of labored breathing, wheezing, coughing, and bronchial constriction, and are a major cause of disability resulting in poor quality of life and high health care costs ^2^. To mitigate exacerbations caused by IgE, anti-IgE therapeutics such as omalizumab block the binding of IgE to receptors ^3^. However, response to anti-IgE therapeutics is inconsistent and depends on multiple factors, including periostin, eosinophil levels in serum ^4^ and allergic sensitization ^5,6^. Therefore, investigating the molecular underpinnings associated with higher serum total IgE may help illuminate biological and chemical pathways associated with clinical response, ultimately informing the best treatment approaches.

To this end, several single-omic studies have elucidated genes and metabolites associated with total IgE levels in both children and adults. These studies implicated, among others, the metabolites valine and tyrosine ^7^, Interleukin 13 ^8,9^, inflammatory and defense response, and cytokine pathways _10_, and leukocyte, lymphocyte, and mononuclear cell proliferation ^11^, all of which are broadly associated with primarily a T helper 2 (Th2) inflammatory response.

However, the understanding of pathways associated with total IgE levels in asthma is still incomplete given the context of high inter-individual heterogeneity in demographics, exposure, and genetics ^2,12^. Notably, the transcriptome and metabolome capture complementary information relevant to these sources of heterogeneity ^13^. Thus, integrating the metabolome with the transcriptome should yield a more holistic picture of the complex metabolic processes underlying allergy responses and total IgE levels in pediatric asthma across heterogeneous groups. However, no studies have evaluated the metabolome and transcriptome in tandem in the context of total IgE. In this study, we evaluated the interplay between the plasma metabolome and transcriptome as it relates to total IgE levels among children with asthma from two independent cohorts: 1) the Childhood Asthma Management Program (CAMP) cohort ^14^ (N=564); and 2) the Genetic Epidemiology of Asthma in Costa Rica Study (GACRS) ^15^ (N=309). In each cohort, total IgE-dependent gene-metabolite associations were identified using a linear modeling approach that included an interaction term between gene expression and serum total IgE and adjusted for clinical covariates ^16,17^. Associations were validated using a null (data permutation) model, and pathway and chemical class enrichment analyses were performed using a comprehensive resource of metabolite and gene annotations ^18,19^. Analyses revealed total IgE-dependent gene-metabolite associations shared between the two cohorts that underlie key metabolic processes relevant to allergic and inflammatory responses. These include the glycine, serine, and threonine metabolism pathway, phospholipid pathways, and possible crosstalk between these pathways and Interleukin-1 signaling.

## Materials and Methods

### The Childhood Asthma Management Program (CAMP)

CAMP ^14^, available from ClinicalTrials.gov register NCT00000575, was a multi-center, randomized, double-masked, clinical trial designed to determine the long-term effects of inhaled treatments for mild to moderate asthma in children. From December 1993 to September 1995, CAMP recruited 1,041 children aged 5 to 12 years at baseline with mild to moderate asthma from 8 sites in North America ^20^. Treatments were randomized to either nedocromil, budesonide, or the placebo arm, and then followed up over the 5 to 6 years of the trial period. All children completed a protocol which included questionnaires and collection of blood. A follow-up study to the primary trial included age, sex, height, and weight and extracted blood samples from 620 CAMP subjects at early adulthood (after trial completion) for gene expression profiling when asthma was not exacerbated, with an average age of 16 years at follow-up ^21^.

### The Genetic Epidemiology of Asthma in Costa Rica Study (GACRS)

GACRS ^15^ recruited a total of 1,165 children aged 6-14 years with mild to moderate asthma from 140 schools across the Central Valley of Costa Rica between February 2001 and August 2008. At enrollment, all children completed a protocol similar to that of the children in CAMP, including blood collection and quantification of serum total IgE at the same timepoint. The questionnaire was a translated version (from English to Spanish) of the one used in the Collaborative Study on the Genetics of Asthma ^22^. Unlike CAMP, GACRS was a cross-sectional observational study. Written parental and participating child consent was obtained. The study was approved by the Partners Human Research Committee at Brigham and Women’s Hospital (Boston, USA) (Partners Human Research Committee; Protocol#: 2000-P-001130/55) and the Hospital Nacional de Niños (San José, Costa Rica).

### Data Collection

The metabolomic samples used in our analysis were collected from plasma in 953 children and the transcriptomic samples from whole blood in 609 children during the follow-up timepoint 4 years later. Serum total IgE level was determined using the UniCAP 250 system and converted to a log_10_ scale for analysis. The CAMP study was approved by the Institutional Review Board of Partners Healthcare (Partners Human Research Committee; Protocol#: 1999-P-001549/29), by all 8 CAMP clinical centers and by the CAMP Data Coordinating Center. Each child’s parent or guardian signed a consent statement, and the clinics also obtained each child’s assent.

### Metabolomic Profiling

Plasma metabolomic profiling was conducted by the Broad Institute using 4 complementary liquid chromatography mass spectrometry (LC-MS) platforms as part of the Trans-Omics for Precision Medicine (TOPMed) initiative ^23^, i.e., Reversed-Phase C8 Chromatography/Positive Ion Mode (C8-pos), Reversed-Phase C18 Chromatography/Negative Ion Mode (C18-neg), Hydrophilic Interaction Liquid Chromatography/Positive Ion Mode (HILIC-pos), and Targeted Negative Ion Mode (Amide-neg) ^24,25^ in 953 children in CAMP and 1,155 children in GACRS. Data processing and quality control (QC) was performed using methods previously described by Kelly et al ^26^. Briefly, metabolite features with undetectable/missing levels for >75% of study samples or a coefficient of variance within QC samples greater than 25% were excluded. All missing values were imputed using the *k*-nearest-neighbor imputation method with *k* = 3. To confirm IDs during analytical runs, authentic standards as well as pooled QC samples were analyzed. Metabolites were analyzed as measured LC-MS peak areas, and log10-transformed and unit-scaled prior to analysis. After these QC and scaling steps, we filtered out metabolites with > 20% minimum-imputed values across samples, resulting in 3,756 metabolites in the GACRS cohort and 3,742 in the CAMP cohort, of which 583 named metabolites were present in both cohorts. Named metabolites with variance at or below the 5^th^ percentile were removed. A total of 553 metabolites passed this threshold in the GACRS cohort and 545 in the CAMP cohort, 529 of which were in common and used in our statistical analyses.

### Transcriptomic Profiling

Whole-blood gene expression profiles were generated with 47,009 probes from the Illumina HumanHT-12 v4 Expression BeadChip, all of which passed stringent and commonly used quality control (QC) metrics (i.e., removal of failed arrays, probes with low outlying log_2_ intensities (<5), and probes with poor signal-to-noise ratios (95^th^ percentile / 5^th^ percentile) ^27^. Expression data were log_2_-transformed and quantile-normalized as a single batch using the *lumiT* and *lumiN* functions, respectively, from the R package *lumi* (version 2.22) ^28,29^. A standard non-specific variance filter was applied to the expression data using the *nsFilter* function from the R package *genefilter* (version 1.52) ^30^. Data were collapsed to a single probe per gene based on the largest interquartile range of expression variance. Genes not annotated with a valid Entrez gene identifier or Human Genome Organization (HUGO) gene symbol or with variance at or below the 5^th^ percentile were removed. A total of 23,722 genes passed this threshold in the GACRS cohort and 23,723 in the CAMP cohort, 22,772 of which were in common and included in our statistical analyses.

### Data Pretreatment

**Supplementary Figure 1** illustrates the data pretreatment procedure. Only samples that could be run on all platforms and that included serum total IgE levels, weight, height, age, race/ethnicity (CAMP only) and study arm (CAMP only) as well as metabolomic and transcriptomic data were retained. In the CAMP clinical data, a single variable combined race and ethnicity (i.e., “Hispanic” was represented as a racial group). Race/ethnicity was omitted from GACRS analyses because the study population was ethnically homogeneous. Levels of 2-deoxyuridine, reduced glutathione, and indole-3-propanoic acid had variances in GACRS that exceeded the maximum variance of any metabolite in CAMP. Upon further inspection, these outlier values were attributed to 10 samples in GACRS, which were removed as they did not pass the generalized extreme Studentized deviate (ESD) test for outliers with α = 0.0001. In total, 309 GACRS and 564 CAMP samples were analyzed.

### Identification of Gene-Metabolite Relationships Dependent on Total IgE

Pairwise linear models were used to capture IgE-dependent gene-metabolite relationships using Integration of Omics Data Using Linear Modeling (IntLIM) 2.0 ^17^, described in Equations 1-2 for GACRS and CAMP, respectively.

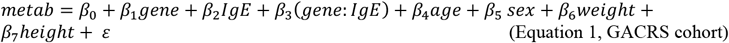

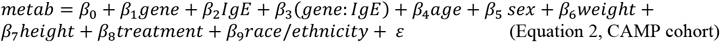

The significance of the statistical interaction between gene expression and serum IgE *(β*_*3*_) denotes the strength of the total IgE-dependent gene-metabolite relationship. All models are adjusted for age, sex, weight, height, and for CAMP only race/ethnicity and treatment arm. The *P*-value of the *β*_*3*_ coefficients were corrected for multiple comparisons (Benjamini-Hochberg False Discovery Rate (FDR)) ^31^. Models with significant FDR-adjusted *P*-values (< 0.05), *β*_3_ coefficients in > 80^th^ percentile; and model *R*^*2*^ > 0.1 were considered statistically significant in each cohort.

### Model Validation

An additional model validation, based on randomly permutated data, was performed using IntLIM 2.0 to ensure that statistically significant IgE-dependent gene-metabolite pairs identified within each cohort were robust when validated against a null model (e.g., not uncovered in permuted data). To do this, serum IgE was randomly assigned to samples in 100 individual permutations while the levels of genes, metabolites, and other covariates retained their original assignment. For each permutation, IgE-dependent gene-metabolite association models were applied in the same manner as the original unpermuted data. Associations present in fewer than 10 random permutations (out of 100) were considered non-random and retained as significant.

### Functional Analyses

For each cohort, a network was constructed, where metabolites and genes were nodes, and IgE-dependent gene-metabolite associations were the edges. Metabolite hubs were defined as metabolites with at least 10 edges (i.e., having IgE-dependent associations with at least 10 genes). Pathway, metabolite chemical class, and reaction-based functional analyses were carried out using Relational Database of Metabolomic Pathways (RaMP-DB) 2.0 ^32^. For each cohort, metabolites and genes from significant IgE-dependent associations were utilized as input pathway and chemical class enrichment. Fisher’s tests were calculated for enrichment analyses. We further evaluated each significant gene-metabolite pair in CAMP and GACRS to determine whether it corresponded to a known chemical reaction. Information regarding each reaction was extracted from the RaMP-DB reaction label for Rhea ^33^ reactions. For the Human Metabolome Database (HMDB) ^34^, the protein corresponding to the gene in the gene-metabolite pair was found using manual lookup on the National Center for Biotechnology Information website . The protein and metabolite were then used to further query the form of the reaction in HMDB.

## Results

Two independent cohorts of children with mild to severe asthma, CAMP (N=564) and GACRS (N=309), were utilized in this study. **Table 1** provides a patient summary for both cohorts after QC and data processing. Notably, serum total IgE value distributions were similar between the two cohorts (**Supplementary Figure 2**). We note that height, and weight differed significantly by *t*-test which was likely driven by the fact that the CAMP population was significantly older at blood collection (*P-*value < 2.2e-16 for all).

**Table 1.**
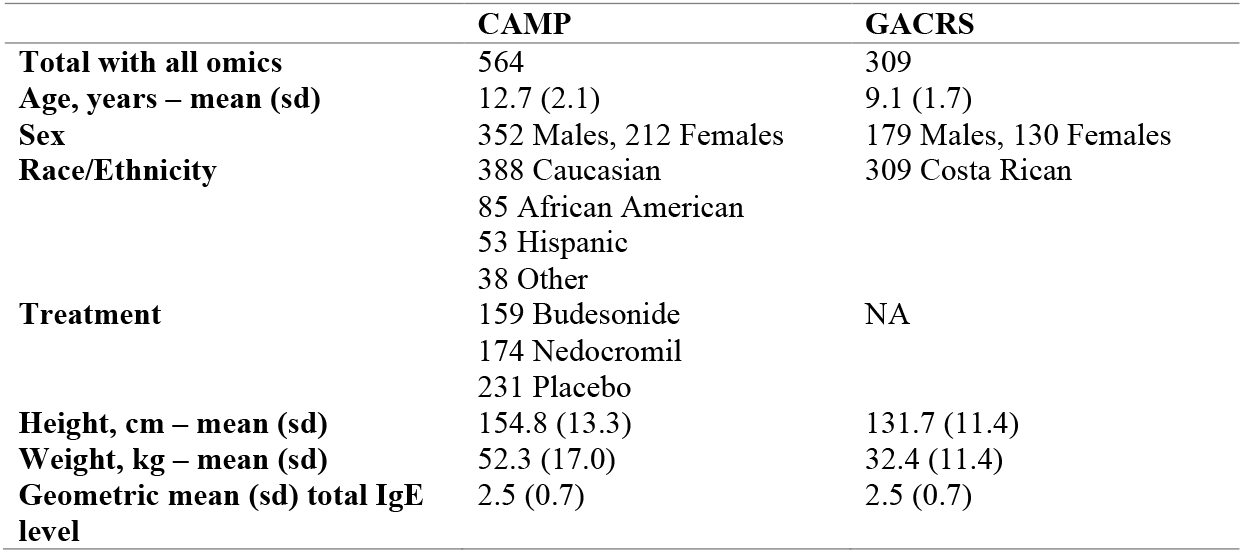
Characteristics of the CAMP and GACRS cohort participants.

Figure 1. illustrates our study workflow. Plasma metabolomic and transcriptomic profiles from the same samples were generated in both cohorts, resulting in 22,772 gene and 529 named metabolite measurements in each cohort. The measured metabolites consisted primarily of lipids and lipid-like molecules, especially glycerophospholipids (GPL), glycerolipids, and fatty acyls; however, other represented classes of molecules included organic acids and derivatives and organoheterocyclic compounds (**Supplementary Figure 3**). IgE-dependent gene-metabolite associations were elucidated using IntLIM 2.0 ^17^, which returns coefficients that describe the statistical interaction of genes and IgE levels (see Methods). A positive coefficient implies that the relationship between a gene and metabolite increases as IgE levels increase. Metabolites and genes from statistically significant associations were then subject to functional analyses to identify enriched pathways and chemical classes. Further, we evaluated whether significant metabolite-gene associations were supported by known reactions.

**Figure 1.**
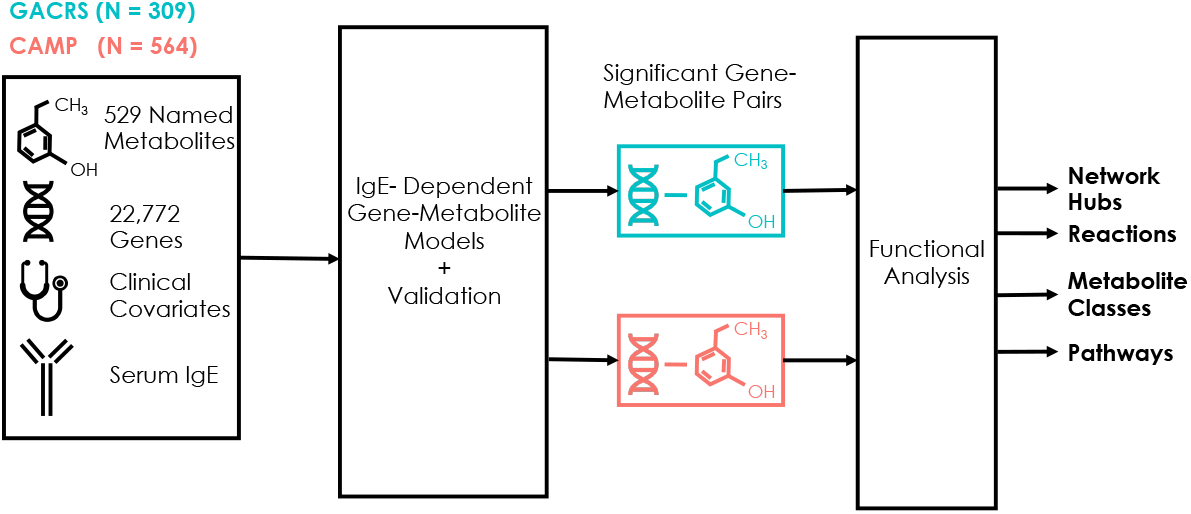
Study workflow. Measurements of 22,772 genes and 529 named metabolites from two independent cohorts, GACRS and CAMP, were used to identify total IgE-dependent gene-metabolite associations (|*β*_*3*_|_*perc*_ > 0.8, *p* < 0.05, *R*^*2*^ > 0.1). To help interpret these associations, pathway and chemical enrichment analyses were performed on metabolites and genes from these associations and reactions that include both the gene and metabolite in an association were identified.

### Total IgE-Dependent Gene-Metabolite Associations in GACRS and CAMP Highlight Role of Guanidinoacetic Acid

For both GACRS and the follow-up timepoint in CAMP, all possible gene-metabolite pairs (12,046,388 total combinations of 529 metabolites and 22,772 genes) were evaluated to identify those that were IgE-dependent. After filtering (|*β*_*3*_|_*perc*_ > 0.8, *p* < 0.05, *R*^*2*^ > 0.1), 29,885 pairs were significant in CAMP, and 1,617 pairs were significant in GACRS after permutation validation **(Supplementary Tables 1, 2, and 3)**. While the overall range of *R*^*2*^ and the distribution of *β*_*3*_ did not differ considerably between GACRS and CAMP, CAMP had more significant *P*-values overall **(Supplementary Figures 4-7)**, which may be due to CAMP having a higher sample size than GACRS. The similarity in *R*^*2*^ range implies that the learned models fit the data equally well in both cohorts.

From the significant pairs, we identified 136 CAMP and 72 GACRS metabolites involved in multiple IgE-dependent gene associations. Metabolite hubs, defined as metabolites associated with more than 10 genes in both cohorts, are listed in **Table 2**. The hubs identified are glycine, diacetylspermine, guanidinoacetic acid (GAA), Lysophosphatidylcholine (LPC)(18:1), pregnenolone sulfate, and Phosphatidylcholine (PC)(P-34:1)/(O-34:2) and are likely to be involved in determining serum total IgE.

**Table 2.** Number of significant IgE-dependent associations found in the 6 metabolite hubs, defined as metabolites associated with > 10 genes in both CAMP and GACRS.

| Metabolite Name | Number of Associations in<br>*CAMP* | Number of Associations in<br>*GACRS* |
| --- | --- | --- |
| Glycine | 573 | 43 |
| Diacetylspermine | 385 | 100 |
| Guanidinoacetic acid<br>(GAA) | 333 | 283 |
| LPC(18:1) | 145 | 19 |
| Pregnenolone sulfate | 75 | 135 |
| PC(P-34:1)/PC(O-34:2) | 27 | 32 |

Further, consistency between the cohorts was observed at the individual gene-metabolite pair level and the individual genes and metabolites contained therein. Specifically, 9 gene-metabolite pairs had IgE-dependent associations in the same direction in both cohorts (**Table 3**).

**Table 3.** Statistically significant gene-metabolite pairs shared between CAMP and GACRS cohorts. * *β –* coefficient of the statistical interaction between gene expression level and serum IgE. CI – confidence interval.

| *Metabolite* | *Gene* | *CAMP* |  | *GACRS* |  |
| --- | --- | --- | --- | --- | --- |
| | | Interaction $\beta$<br>(95% CI) | FDR | Interaction $\beta$<br>(95% CI) | FDR |
| **GAA** | PTPN12 | 0.06 (0.03, 0.08) | <0.01 | 0.08 (0.04, 0.11) | 0.04 |
|  | HNRNPH2 | 0.06 (0.03, 0.08) | <0.01 | 0.09 (0.05, 0.13) | 0.04 |
|  | PELI1 | 0.05 (0.03, 0.07) | <0.01 | 0.08 (0.04, 0.12) | 0.03 |
|  | CHMP1B | 0.05 (0.03, 0.08) | <0.01 | 0.08 (0.04, 0.12) | 0.03 |
|  | C9orf6 | 0.06 (0.02, 0.09) | 0.04 | 0.12 (0.07, 0.18) | 0.03 |
|  | RCHY1 | 0.07 (0.02, 0.11) | 0.05 | 0.13 (0.07, 0.18) | 0.02 |
|  | VANGL2 | 0.08 (0.03, 0.12) | 0.05 | 0.17 (0.10, 0.24) | 0.02 |
| **Hydroxyproline** | MGC52282 | 0.06 (0.03, 0.09) | 0.01 | 0.08 (0.04, 0.12) | 0.04 |
| **Diacetylspermine** | DNAJC11 | -0.09 (-0.15, -0.04) | 0.04 | -0.27 (-0.39, -0.16) | 0.01 |

Notably, GAA is involved in 7 of these 9 IgE-dependent gene-metabolite associations. The genes associated with GAA are involved in Interleukin-1 signaling (PTPN12 and PELI1), mRNA processing and splicing (HNRNPH2), and DNA damage bypass (RCHY1). Because all these pathways are involved in inflammatory response, the associations suggest possible crosstalk between GAA and inflammatory response. Lastly, we note that a total of 770 genes and 50 metabolites involved in IgE-dependent associations were shared between both GACRS and CAMP. For comparison, 12,221 genes and 172 metabolites were involved in IgE-dependent associations in CAMP while 1,283 genes and 123 metabolites were involved in IgE-dependent associations in GACRS, respectively.

### Functional Analysis Reveals Perturbation of Glycine, Serine, and Arginine Metabolism in Serum IgE-dependent Gene-Metabolite Pairs

Significant IgE-dependent pairs in CAMP (29,885) comprise 47 HMBD reactions and 4 Rhea reactions, while those significant in GACRS represented a single HMDB reaction, i.e., each reaction includes both the gene and the metabolite from a significant gene-metabolite pair **(Supplementary Tables 4-5)**. The reaction represented in GACRS pairs (1,617) was the conversion of L-Arginine and glycine into ornithine and GAA, catalyzed by glycine amidinotransferase (GATM). Notably, this reaction was also significant (|*β*_*3*_|_*perc*_ > 0.8, *p* < 0.05, *R*^*2*^ > 0.1) in CAMP but was removed after the permutation testing. Other CAMP reactions related to this single GACRS reaction included conversion of glycine to serine and two reactions belonging to the citric acid cycle, which is downstream of glycine to serine conversion in the glycine, serine, and threonine metabolism and of the arginine and proline metabolism pathways. These reactions included conversion of Adenosine Triphosphate (ATP) to Adenosine Diphosphate (ADP) via creatine and conversion of Acyl Coenzyme A to Coenzyme A. We also note that the conversion of glycine to serine was borderline significant in GACRS (FDR = 0.13), suggesting that it may also be perturbed with serum IgE level in both cohorts.

#### Pathways

Pathways that included the 12,221 genes and 172 metabolites from significant total IgE-dependent gene-metabolite pairs in CAMP and the 1,283 genes and 123 metabolites in GACRS in both cohorts were also evaluated, with all measured genes and metabolites included as the background. While none of the pathways achieved statistical significance (Benjamini-Hochberg-adjusted *P*-value < 0.05), we found that the top 50 pathways (sorted by nominal *P*-value) represented in each cohort included 29 shared pathways **(Figure 2). Supplementary Tables 6-7** include all pathways. Shared pathways included glycine metabolism, creatine pathways, serine metabolism, and amino acid metabolism, further supporting the relationship between glycine and serine metabolism and serum IgE level. Moreover, shared glutathione, creatine, and proline pathways support the relationship between arginine and proline metabolism and serum IgE level.

**Figure 2.**
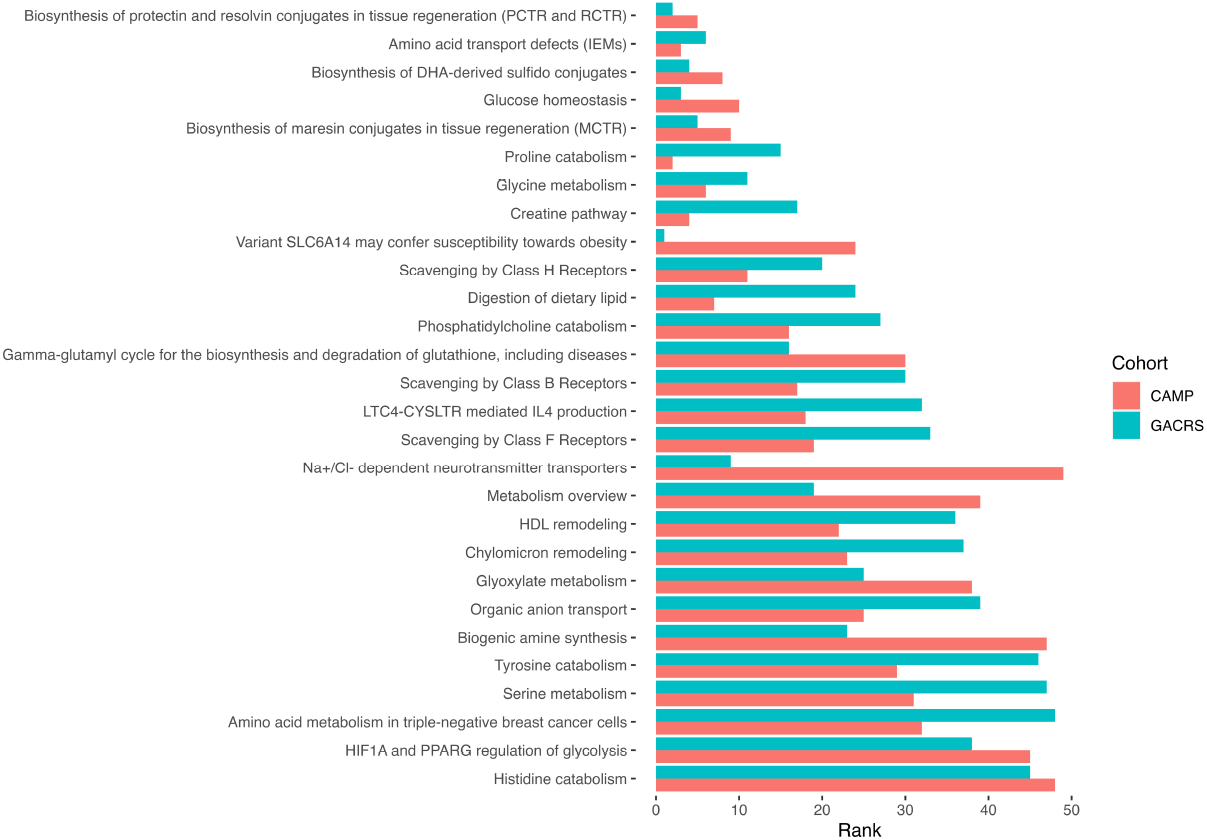
Top 50 pathways, sorted by enrichment *P*-value rank, represented in the GACRS and CAMP IgE-dependent gene-metabolite pairs.

#### Chemical Classes

We further performed class enrichment analysis to provide an overview of the chemical classes of metabolites represented in total IgE-specific gene-metabolite pairs. We found that the significant associations in CAMP were enriched for glycerophospholipids and glycerolipids with Benjamini-Hochberg adjusted *P*-value < 0.05 **(Supplementary Table 8)**. The significant associations in GACRS were enriched for lipids and lipid-like molecules, with glycerolipids significant prior to adjustment **(Supplementary Table 9)**. Notably, glycerophospholipid metabolism is a downstream pathway of glycine, serine, and threonine metabolism that also overlaps with glycerolipid metabolism, suggesting dysregulation of the glycerophospholipid and glycerolipid pathways via glycine, serine, and threonine metabolism could be involved in modulation of total IgE levels.

## Discussion

Through integration of metabolomic and transcriptomic profiles, we uncovered associations that implicate a potential role of GAA and glycine in modulating serum total IgE levels of children with asthma. Prior literature shows that with increased GAA (supported by the positive *β*_*3*_ values), the levels of arginine needed for creatine synthesis decrease, freeing arginine for other functions (such as its role in reducing inflammation) ^35^. This presents a possible indirect mechanism by which GAA influences total IgE levels. Additionally, mitochondrial, inflammatory and DNA damage pathways were represented in our uncovered IgE-dependent gene-metabolite pairs, which may reflect activity of creatine (derived from GAA) in inflammatory response via mitochondrial dysfunction and DNA damage. The role of creatine in these processes has previously been studied outside of the context of asthma ^36-40^.

Associations shared between the two cohorts reflected reactions participating in or downstream of pathways in which GAA and glycine are integral: namely, (1) amino acid metabolism, (2) glycine, serine, and threonine metabolism, and (3) arginine and proline metabolism. Specifically, the conversion of L-arginine and glycine into ornithine and GAA as represented in GACRS is central to glycine, serine, and threonine metabolism as well as arginine and proline metabolism, and the conversion of glycine to serine as represented in CAMP is central to glycine, serine, and threonine metabolism. The Tricarboxylic Acid (TCA) cycle reactions represented in CAMP are downstream of all three pathways. Moreover, amino acid, glycine, serine, creatine, glutathione, and proline pathways were shared between both cohorts, and the glycerophospholipid class was enriched in CAMP. These findings uncover a possible mechanistic basis for the total IgE levels observed in children with asthma via GAA and its pathways, and thus provide opportunities for targeting these mechanisms.

The relationship between these relevant pathways and total IgE level is consistent with previous work showing that that glycine and serine reduce inflammation ^41^ and that levels of creatine and glycine in plasma decrease after inhalation of budesonide and salbutamol by children with asthma _42_, indicating increased glycine and serine metabolism. Further, amino acids have previously been implicated in asthma status and lung function ^43,44^, and serine protease inhibitors have been shown to mitigate inflammatory cytokine production and to suppress pro-inflammatory genes in a mouse model of asthma ^45^. Finally, l-Arginine supplementation has been shown to inhibit nitric oxide producing enzymes (associated with inflammation) via the NOS and arginase pathways in animal and cell culture models ^46,47^. L-Arginine supplementation also decreases asthma exacerbations in a subset of clinical trial subjects driven by metabolites that include creatine, taurine, linoleic acid, and α-Glutamylcysteine (Glu-Cys) ^48^.

The strengths of our analyses include the following: (1) we integrated metabolomics and transcriptomics data in an IgE-dependent context in pediatric asthma, which have not been investigated previously, (2) transcriptomic and metabolomic measurements were performed on the same blood samples in GACRS and on samples taken 4 years later in CAMP, (3) we replicated the results in two separate cohorts with markedly different clinical and population characteristics, and

(4) we evaluated relevant biological and chemical pathways, which are complementary and circumnavigate the issue of sparse pathway annotations for metabolites in knowledgebases ^49^. One limitation of our analysis is that it is restricted to linear models and thus we are not detecting non-linear relationships which could be biologically relevant. Further, our analysis is correlative and does not reveal causal relationships between metabolites and transcripts or directionality of pathway and reaction dysregulation with increasing serum IgE level. We also note the lack of statistical significance in pathway enrichment analyses, which is likely due to the high number of input metabolites and genes. Despite these limitations, our identified IgE-dependent gene-metabolite pairs were supported by observed chemical reactions and highlight key metabolites, notably GAA, that could modulate serum IgE levels. These observations at a population level offers support for further mechanistic and validation studies that aim to identify potential therapies for asthma exacerbation.

Our integrative analysis of metabolomic and transcriptomic profiles support the potential involvement of GAA, glycine, and related pathways in modulating serum IgE levels of children

with asthma. Notably, our results were consistent across two diverse and independent cohorts, CAMP and GACRS, cohorts that have stark differences in population characteristics.

## Data Availability

https://topmed.nhlbi.nih.gov/

## Acknowledgments

We thank the NIH Individual Graduate Partnership Program for training and mentoring. Further, we thank the Broad Institute for their correspondence regarding the metabolomics data.

## Author Contribution Statement

EAM and JL-S conceptualized the study; STW, RG, MM, JC, RSK, and JL-S contributed to the design and collection of the TOPMed data described herein. CC performed metabolomic profiling. TE and JKS performed the data pretreatment and statistical analysis. RSK, JL-S, EAM, RM, TE, and JKS conceptualized the analytical workflow. RSK, JL-S, EAM, and TE contributed to data interpretation. JB built a custom version of the RaMP-DB database for the analytical workflow. TE, EAM, RSK, JL-S, and RM drafted the manuscript and contributed to revision of manuscript and editing. All authors had final responsibility for the decision to submit for publication.

