## Supplemental Figures for "Consistent Multi-Omic Relationships Uncover Molecular Basis of Pediatric Asthma IgE Regulation"

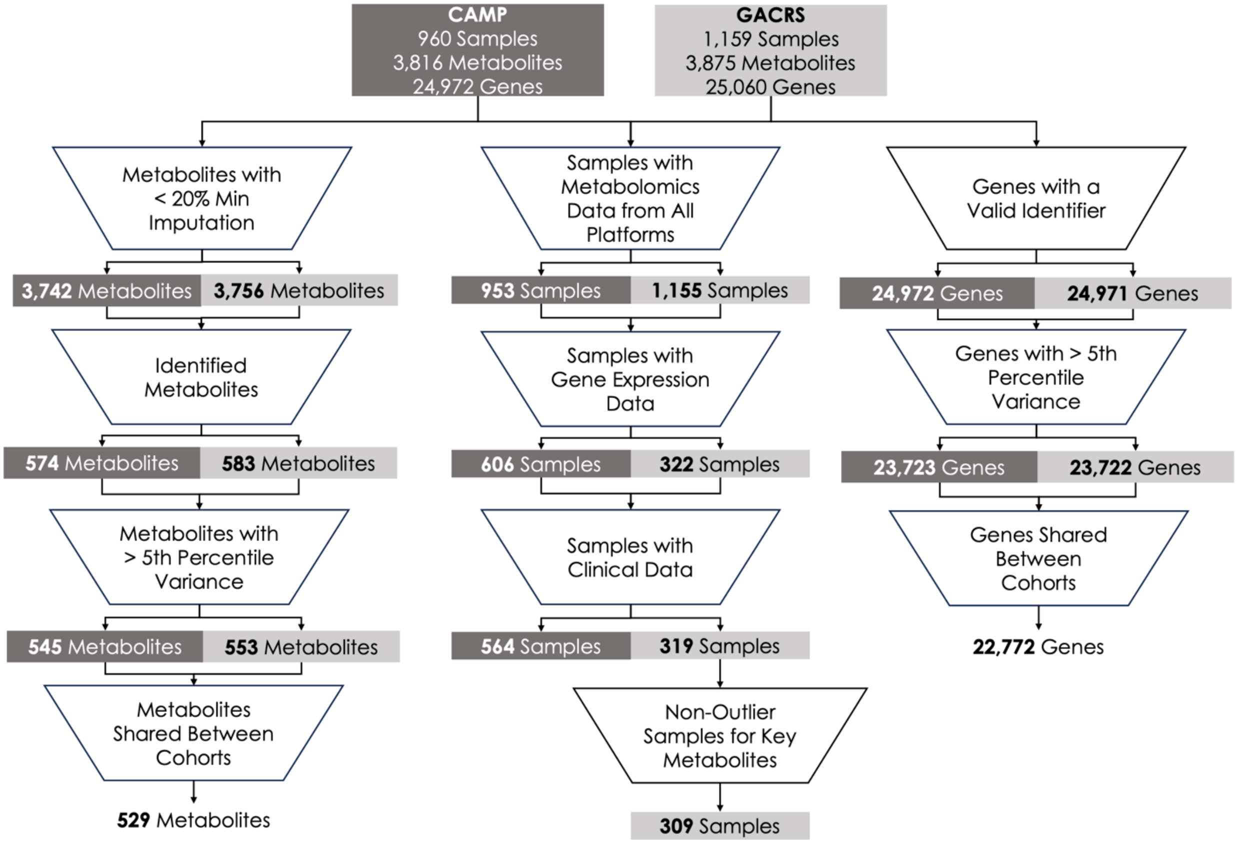


**Supplementary Figure 1:** Data preprocessing workflow applied to the CAMP and GACRS cohorts. A total of 529 metabolites and 22.772 genes were used in our statistical analyses for CAMP (N=564) and GACRS (N=309) cohorts.

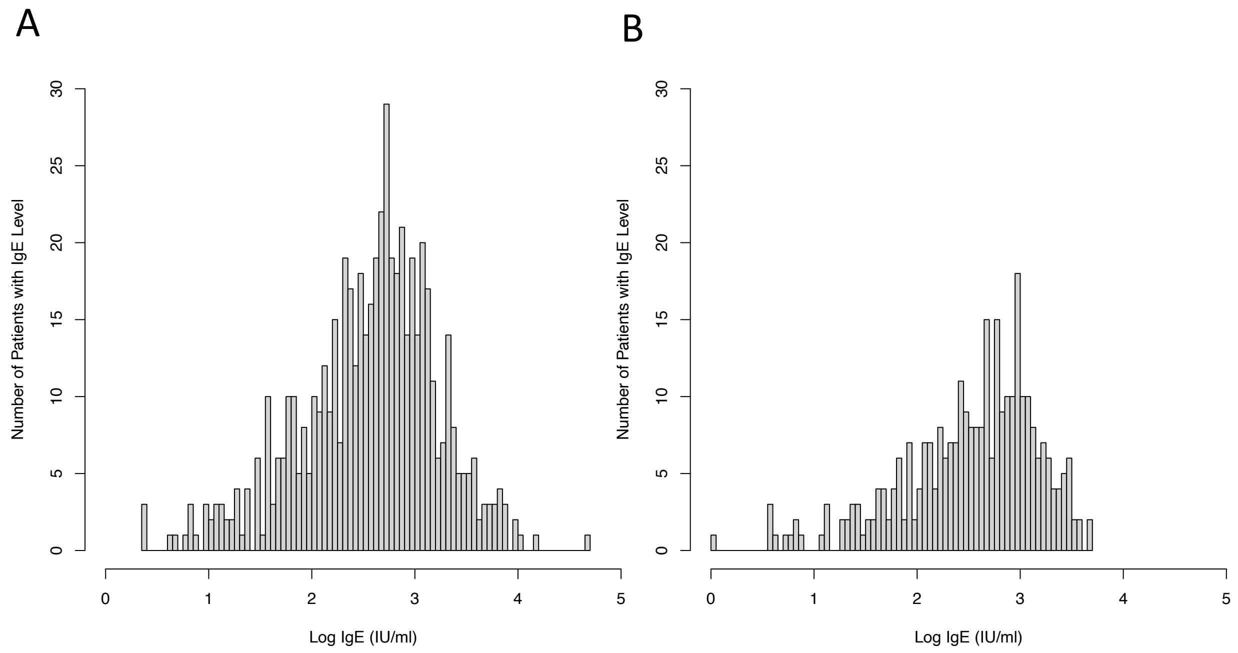


**Supplementary Figure 2:** Histograms of the log(IgE) values for samples in (A) CAMP and (B) GACRS.

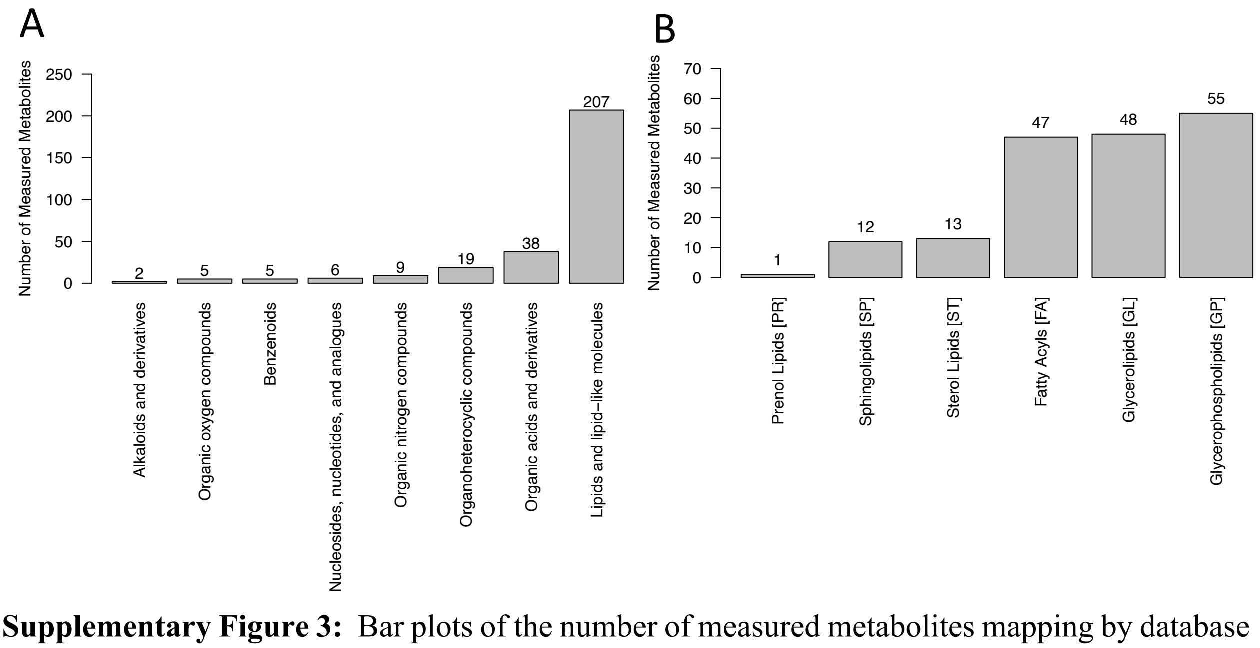


**Supplementary Figure 3:** Bar plots of the number of measured metabolites mapping by database identifier to (A) each ClassyFire Superclass in RaMP 2.0 and (B) each LipidMaps Category in RaMP 2.0.

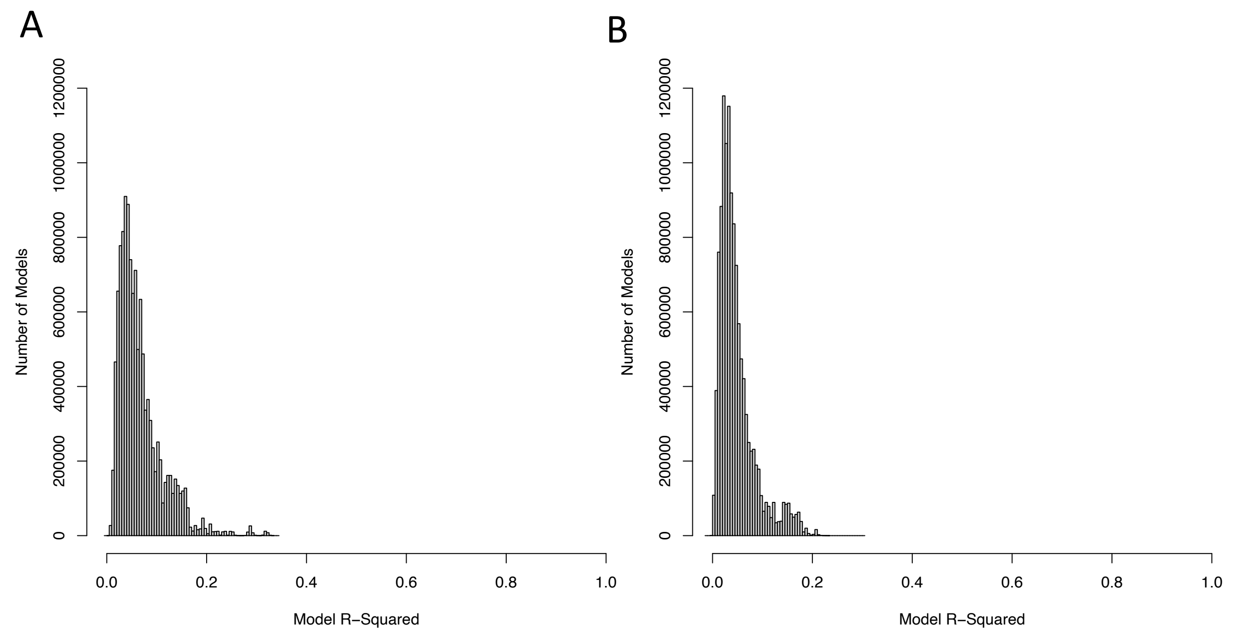


**Supplementary Figure 4:** Histograms of the $R^2$ values for all analyte pairs evaluated in (A) CAMP and (B) GACRS.

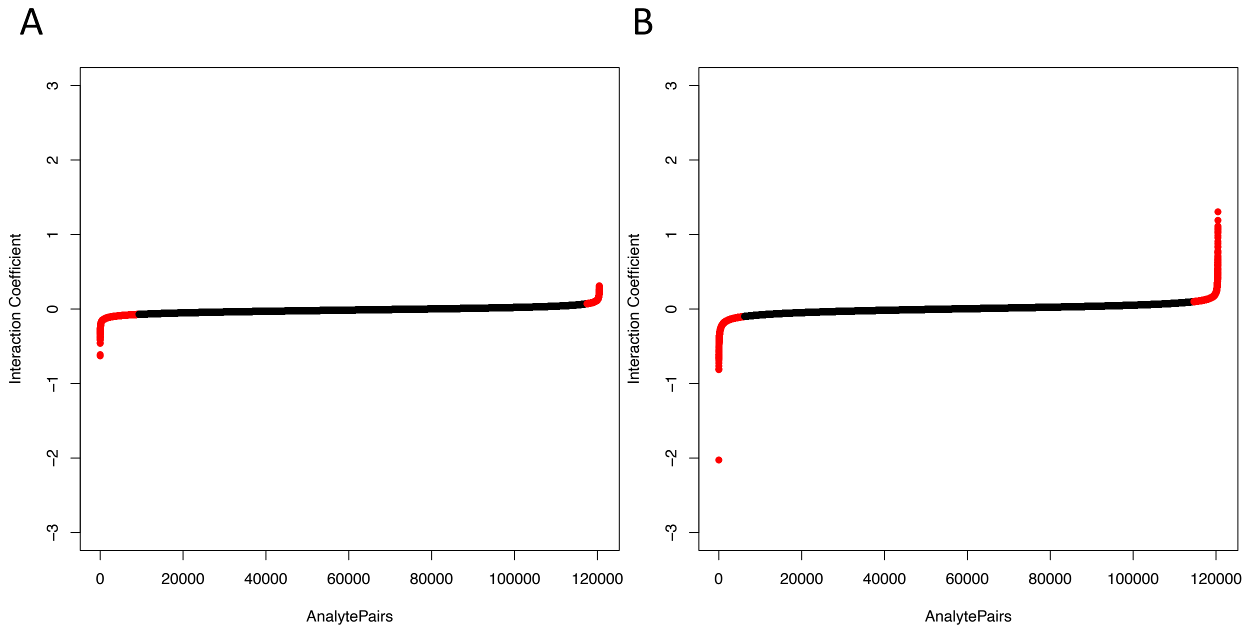


**Supplementary Figure 5:** Plots of the interaction ($\beta_3$) coefficients for all analyte pairs evaluated in (A) CAMP and (B) GACRS. Pairs exceeding the 90th percentile $|\beta_3|$ of are highlighted in red.

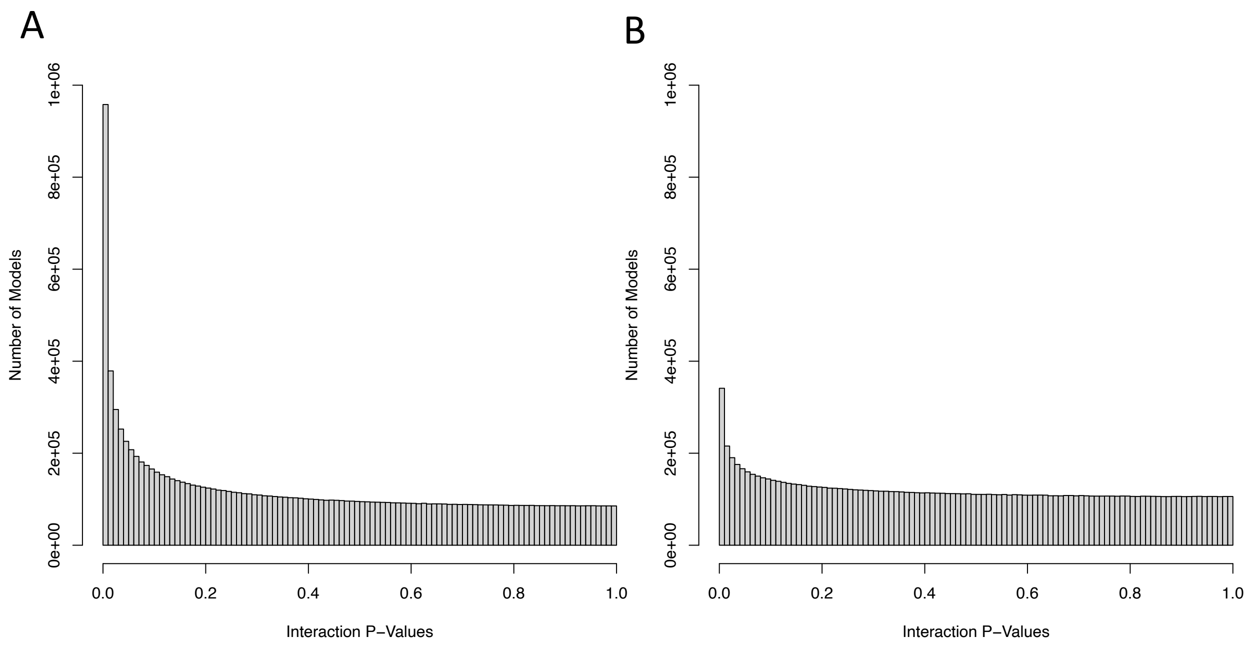


**Supplementary Figure 6:** Histograms of the nominal (unadjusted) p-values for all analyte pairs evaluated in (A) CAMP and (B) GACRS.

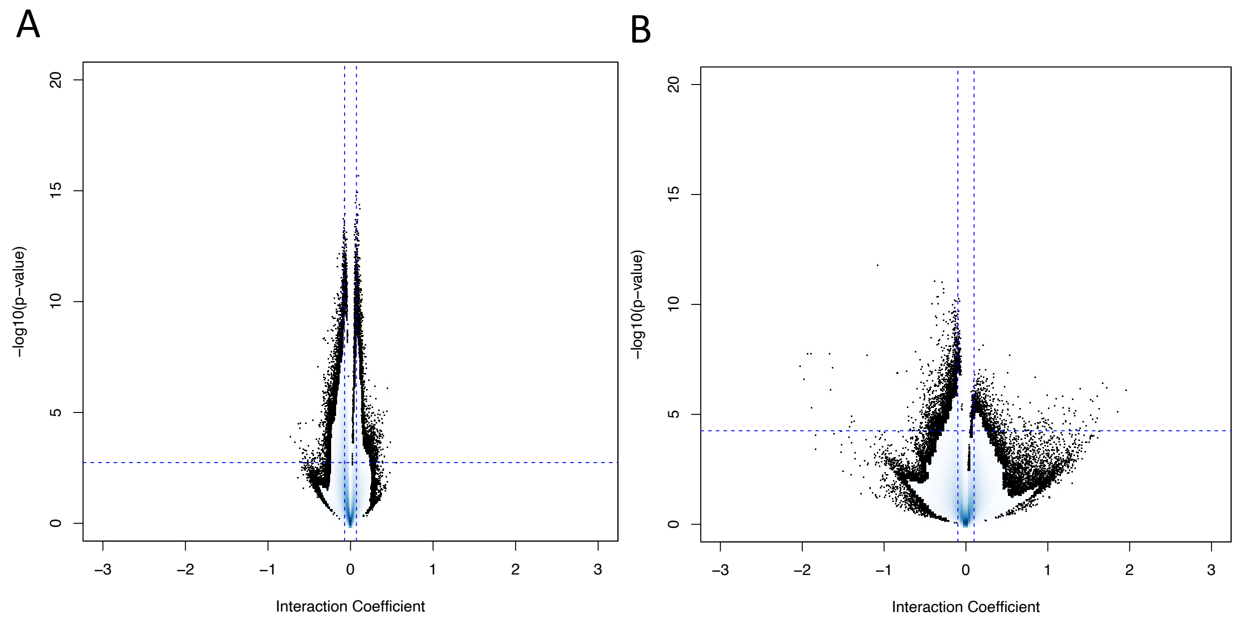


**Supplementary Figure 7:** Volcano plots of the *p*-values and effect sizes (interaction coefficients) for all learned IntLIM models for the (A) CAMP and (B) GACRS cohorts.
